# Assumptions for creating matrices of evidence to estimate overlap of primary studies in overviews of reviews: Protocol for a meta-research study

**DOI:** 10.1101/2023.06.16.23291488

**Authors:** Javier Bracchiglione, Nicolás Meza, Carole Lunny, Dawid Pieper, Eva Madrid, Gerard Urrútia, Xavier Bonfill Cosp

## Abstract

**Introduction:** Overlap of primary studies among systematic reviews (SRs) included in an overview is a major challenge, as it may bias results or artificially increase the precision of the synthesis. Matrices of evidence and corrected covered area (CCA) calculation are recommended methods to manage overlap, but there is little guidance on how to construct these matrices. This research aims to explore variations in the estimation of overlap using CCA matrices under different assumptions.

**Methods:** We will include overviews published in 2023. We will describe the methods used by authors to deal with overlap, and we will calculate a summary CCA (a CCA for the whole matrix of evidence) and a pairwise CCA (a CCA for each possible pair of included SRs), comparing the results under different assumptions that may modify the evidence matrix and thus the CCA. These assumptions include: publication-thread adjustments (i.e. the consideration of each set of references regarding a single primary study as a unique row in a matrix of evidence), scope adjustments (i.e. the consideration only of the SRs and primary studies providing useful data for a given outcome within an overview) and chronological structural missingness adjustments (i.e. the exclusion of primary studies published after a given SR for purposes of CCA calculation). We will assess overlap at an overview and outcome level.

**Discussion:** We propose clear definitions for the key assumptions for creating matrices of evidence. We expect to provide a guide for overview authors to better interpret their CCA estimations.

**Article Summary:** *Strengths and limitations of this study:*

- This protocol explores the assumptions underlying the overlap assessment in overviews of systematic reviews, that so far have not been explicitly addressed.
- These assumptions include scope adjustments, publication-thread adjustments, structural missingness adjustments, and analysis at an overview or outcome level.
- We provide clear definitions for key overlap concepts that will guide authors for making their overlap assessments more explicit when using a matrix of evidence or corrected covered area approach.
- We plan to conduct exploratory analyses under different assumptions in a purposive sample of overview, hence, we will not comprehensively include all the overviews in the study period.
- We will conduct all the analysis calculating the corrected covered area for the whole matrices (overall approach) and for every possible pair of systematic reviews within each matrix (pairwise approach).

## Introduction

Overlap of primary study results among systematic reviews (SRs) is one of the key methodological challenges in the conduct of overviews [1–4]. Proper handling of this issue is crucial, as failure to do so could introduce bias or artificially inflate the precision of the results due to overlap [5]. Overlap can introduce bias if the same study data is included in multiple reviews, as this can inflate the importance of certain studies and skew the overall findings. Additionally, if the studies included in multiple reviews are not representative of the full range of studies on a topic, this can lead to biased conclusions. Overlap can also artificially increase precision, as the same data points may be included multiple times in the analyses. This can overly exaggerate the precision of the results, making them appear more reliable than they actually are.

Matrices of evidence and the calculation of the corrected covered area (CCA) are among the most recommended methods for dealing with overlap [3]. Matrices of evidence are visual grids that show how duplicated primary studies overlap across multiple SRs on the same clinical, public health or policy question, and the CCA is a quantitative approach to evaluate the degree of overlap based on matrices of evidence [6].

Several authors have made recommendations about how to assess and incorporate the calculation of the CCA into overviews [1–4,6–12]. However, these recommendations do not explicitly provide guidance about how to accurately construct a matrix of evidence. Specifically, three aspects of the process of building a matrix have not been clearly defined: (1) Publication-threads adjustment, refers to the practice of treating multiple references related to the same group of patients as a single primary study. This approach corrects the common mistake of considering each reference as a separate study when multiple citations use the same patient data. Publication-threads adjustment considers the whole set of references accounting one unique primary study; (2) scope adjustment, refers to the decision about whether to consider all included primary studies in every SR as rows in the matrix or only those primary studies that provide useful data for the overview (that is, data providing information for an outcome within the overview); and (3) structural missingness adjustment, refers to the decision of whether to consider in the calculation of the CCA those primary studies that were not be included in a given SR (e.g. missing because they were published after the SR, or due to explicit restrictions in the eligibility criteria of a SR such as language exclusion criteria) [9]. Also, a matrix can be generated at an overview level, or authors may present one matrix per outcome. These issues may modify a matrix of evidence, and affect the calculation of the CCA and, therefore, it may impact the overview analysis or interpretation.

The aim of this research is to explore the variations in the estimation of overlap under these three different assumptions for creating a matrix of evidence in the context of overviews.

## Methods

We will conduct a methodological review. We will use the relevant items in the PRISMA guidelines for reporting our research [13,14].

### Eligibility criteria

We will include a purposive sample of overviews published in 2023. We set this chronological threshold since the latest version of the Cochrane handbook for SRs of intervention —which included guidance for conducting overviews— was first published in 2019, and the ‘Preferred Reporting Items for Overviews of Reviews’ (PRIOR) statement was published in 2022 [15,16]. We define an overview as any type of evidence synthesis whose unit of searching, inclusion and data analysis are SRs [5].

### Search methods and selection of studies

We will search the Cochrane Library and MEDLINE/PubMed using previously validated search strategies for retrieving overviews [17,18]. We will restrict our search to articles published between January 1, 2022 and April 1, 2023. Two authors will independently screen articles for eligibility, first by title and abstract, and then by full text. We will solve discrepancies through discussion and consensus.

### Data extraction and analysis

For each included overview, one author will extract the following data: Year of publication, overview scope classification as narrow (one population and one intervention or comparison) or broad (multiple populations or multiple interventions or comparisons) [12], methods used by the authors to deal with overlap according to the MOoR framework (appendix 1) [3], number of included SRs, number of primary studies included in each SR, number of duplicated references for each primary study included in each SR. A second author will cross-check this process. We will not consider ongoing studies as included for matrix purposes. Besides extracting the absolute number of SRs, primary studies and duplicated references of primary studies, we will create matrices of evidence using the GROOVE tool for each included overview [9].

In this protocol, we propose explicit concepts and definitions for the different assumptions that authors may consider when creating a matrix of evidence —and that can, therefore, impact the CCA calculation. These assumptions include:

- Unit of analysis assumption: Matrix construction at the overview or outcome level.
- Assumptions for building the matrix of evidence: The matrix by scope, publication threads, and / or structural missingness.
- CCA calculation approach: Calculate the CCA for the whole matrix (overall approach) and / or for every available pairs of SRs (pairwise approach).

The definitions for each concept and main references are provided in Box 1. For this study, we will first build a ‘default’ matrix of evidence, that is, an overview-level matrix with no scope, publication-thread or structural missingness adjustment, and perform both an overall and pairwise CCA calculation (see Box 1 for definitions).

#### Box 1.

Definition for key overlap concepts

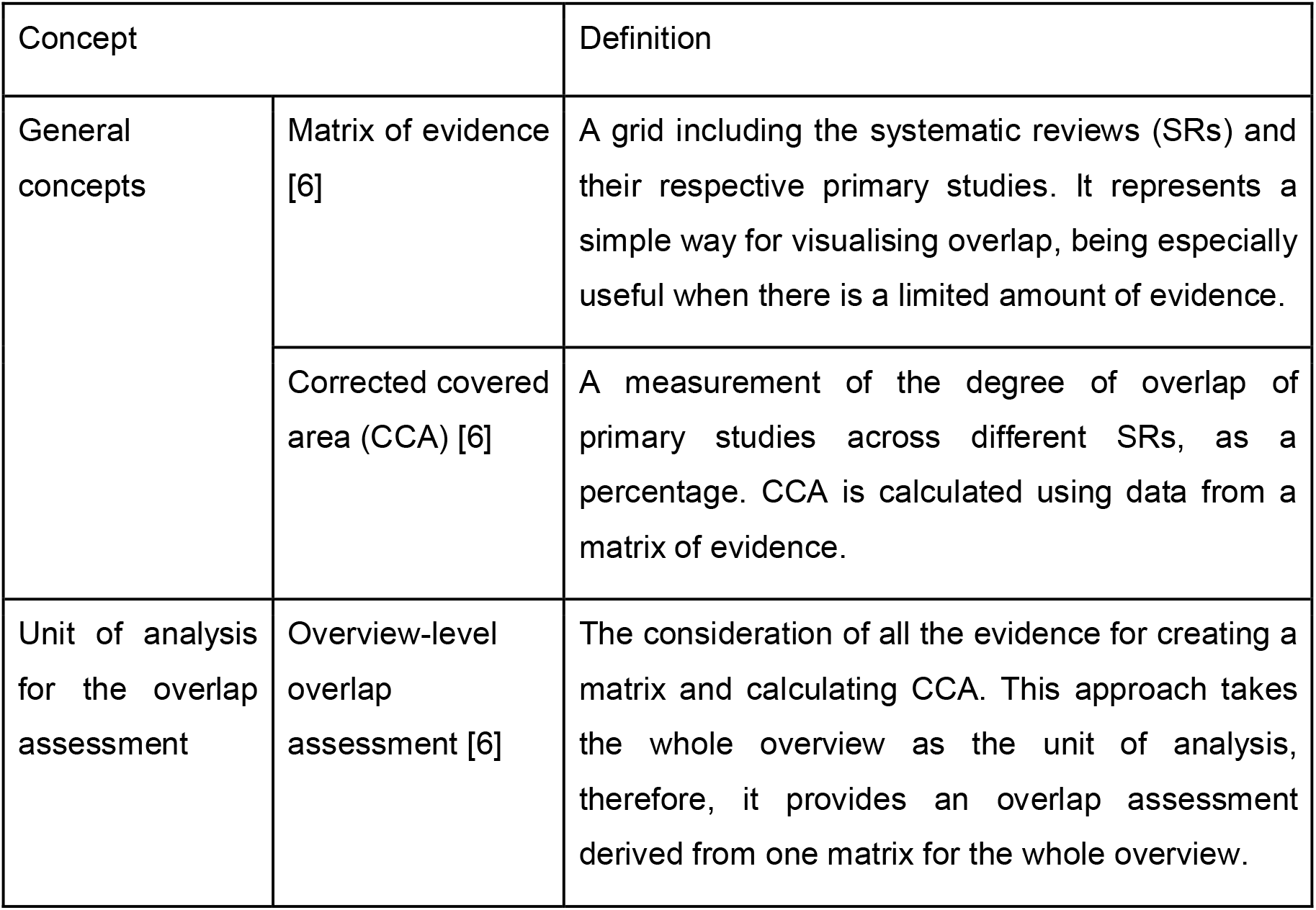
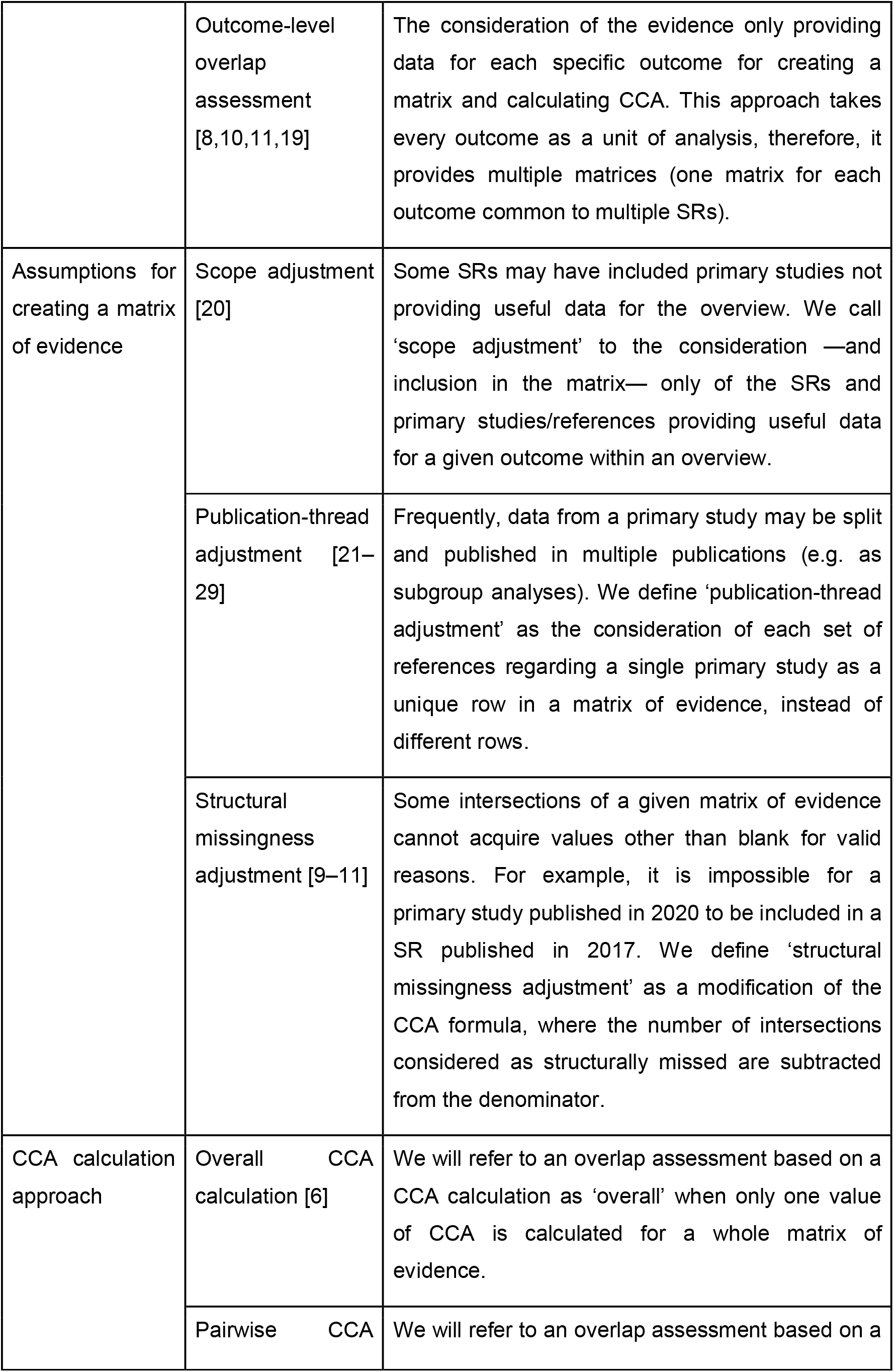
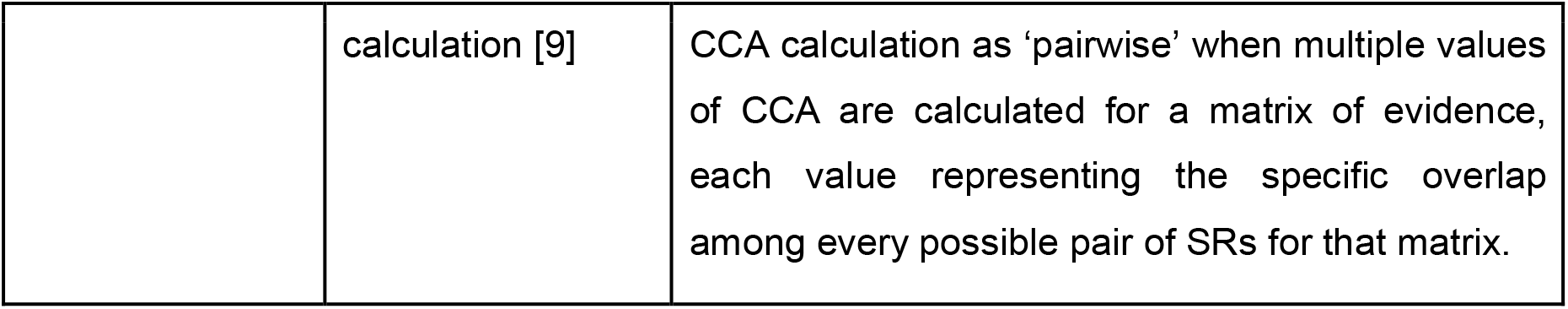

We will then modify this ‘default’ matrix for each overview, creating different scenarios, as described in table 1. In this review, we will define structural missingness to be based exclusively on a chronological basis for adjustment purposes. We will do this by comparing the date of publication of the primary study versus the date of search of a particular SR, considering as a structural missingness any primary study published after the search date of each SR.

**Table 1:** Different scenarios planned for analysis of matrices of evidence and CCA calculations.

|  | Scope adjustment | Publication-thread adjustment | Structural missingness adjustment |
| --- | --- | --- | --- |
| 'Default' scenario | x | x | x |
| Scenario 1 | ✓ | x | x |
| Scenario 2 | x | ✓ | x |
| Scenario 3 | x | x | ✓ |
| Scenario 4 | ✓ | ✓ | x |
| Scenario 5 | ✓ | x | ✓ |
| Scenario 6 | x | ✓ | ✓ |
| Scenario 7 | ✓ | ✓ | ✓ |

We will describe the overall CCA and a pairwise analysis of the CCA for the different scenarios, as provided by the GROOVE tool [9]. We will compare the results of the CCA assessment with the default scenario as reference. All the above-mentioned analyses will be conducted at an overview level. Nevertheless, we will also conduct an analysis at an outcome level using scenario 7, since, theoretically, this would represent an ideal approach (adjusted by scope, publication thread and structural missingness).

## Discussion

CCA calculation is among the most accepted methods for estimating overlap degree among SRs. Since the CCA formula derives from the matrix of evidence, a clear consideration and reporting of the assumptions taken for creating these matrices is necessary for overview authors. In this protocol, we propose clear definitions for the key assumptions for creating different matrices of evidence. We hope these concepts will help authors to improve their methods for conducting and reporting overlap in the conduct of overviews.

Our study will explore how different assumptions for creating matrices of evidence impact the overlap estimation through the CCA. We will make recommendations based on our results. We expect our overlap estimations to be highly heterogeneous under the different scenarios described in table 1. If that is the case, we hope our findings will provide an initial guide for overview authors to better interpret their CCA estimations, and also, encourage future meta-epidemiological research about how the CCA estimations may bias or impact the precision of the findings derived from overviews.

## Data Availability

All data produced in the present work are contained in the manuscript

## Patient and public involvement

No patients were involved in the development of this protocol.

## Ethics and dissemination

No ethical approval is required to conduct this project. We will submit our findings to appropriate scientific conferences, and peer-reviewed journals.

## Acknowledgments

Javier Bracchiglione is a PhD candidate at the Doctorate Program on Biomedical Research and Public Health, Universitat Autònoma de Barcelona, Barcelona, Spain.

## Author contributions

JB developed the study concept and drafted the manuscript of the protocol. NM, CL, DP, EM, GU and XB developed and provided feedback for all sections of the review protocol. Searches of studies and study selection will be performed by JB and NM. Data extraction, analyses and interpretation of the results will be performed by all the authors. All authors reviewed and approved the final version of the manuscript.

## Patient consent for publication

Not required.

## Provenance and peer review

Not commissioned; externally peer-reviewed.

## Data sharing statements

No additional data are available.

## Appendix 1 Methods to address overlap according to the MOoR framework [3]

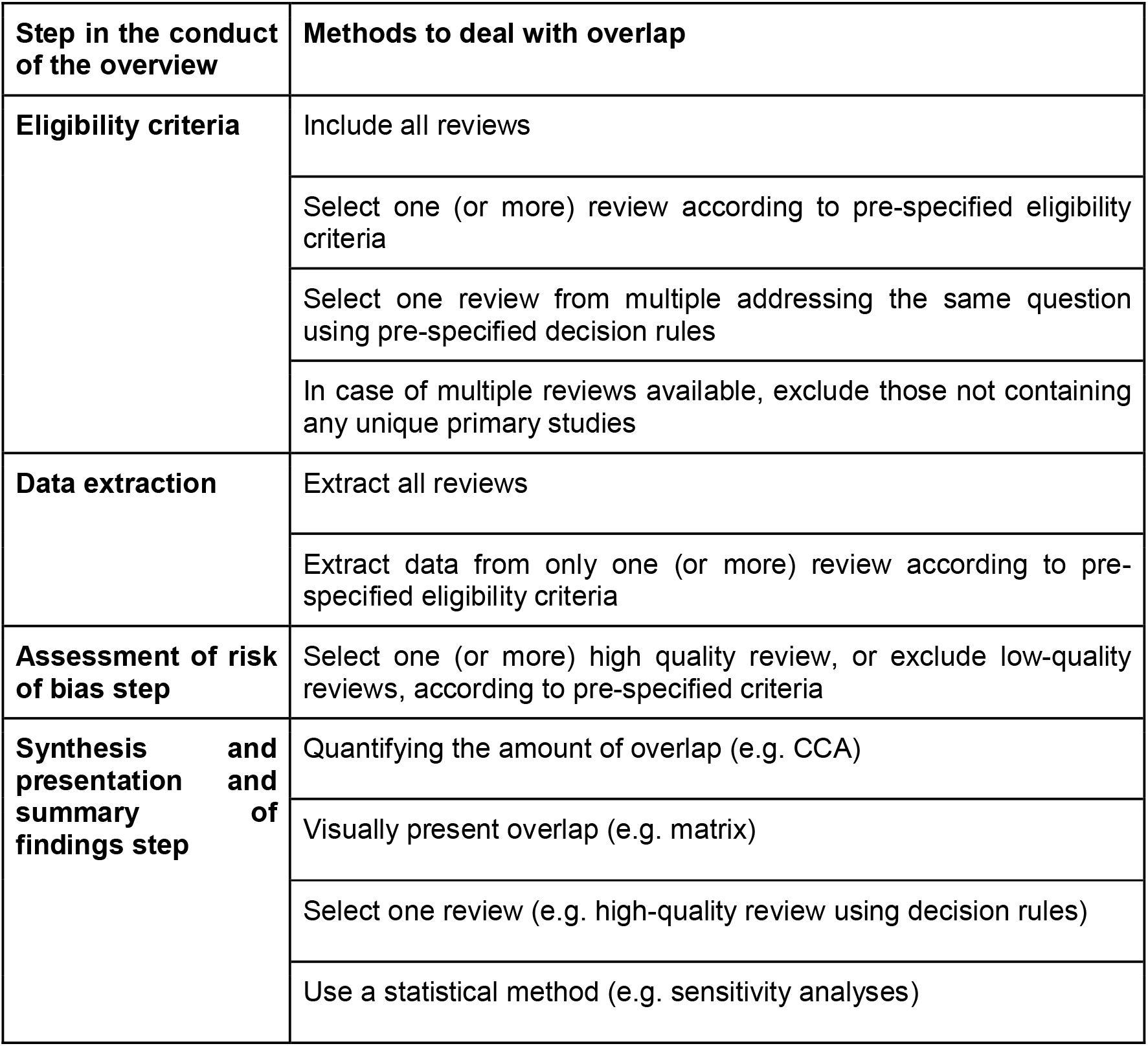

